# Mining for Equitable Health: Assessing the Impact of Missing Data in Electronic Health Records

**DOI:** 10.1101/2022.05.09.22274680

**Authors:** Emily Getzen, Lyle Ungar, Danielle Mowery, Xiaoqian Jiang, Qi Long

## Abstract

Electronic health records (EHRs) are collected as a routine part of healthcare delivery, and have great potential to be utilized to improve patient health outcomes. They contain multiple years of health information to be leveraged for risk prediction, disease detection, and treatment evaluation. However, they do not have standardized formatting, and can present significant analytical challenges– they contain multi-scale data from heterogeneous domains and include both structured and unstructured data. Data for individual patients are collected at irregular time intervals and with varying frequencies. In addition to the analytical challenges, EHRs can reflect inequity– patients belonging to different groups will have differing amounts of data in their health records. Many of these issues can contribute to biased data collection. The consequence is that the data for marginalized groups may be less informative due to more fragmented care, which can be viewed as a type of missing data problem. For EHRs data in this complex form, there is currently no framework for introducing missing values. There has also been little to no work in assessing the impact of missing data in EHRs. In this work, we simulate realistic missing data scenarios in EHRs to adequately assess their impact on predictive modeling. We incorporate the use of a medical knowledge graph to capture dependencies between medical events to create a more realistic missing data framework. In an intensive care unit setting, we found that missing data have greater negative impact on the performance of disease prediction models in groups that tend to have less access to healthcare, or seek less healthcare. We also found that the impact of missing data on disease prediction models is stronger when using the knowledge graph framework to introduce realistic missing values as opposed to random event removal.

## 1 Introduction

In 2009, the Health Information Technology for Economic and Clinical Health (HITECH) act was passed to promote the adoption of health information technology in the United States (Hoerbst and Ammenwerth, 2010). For more than a decade, the use of Electronic Health Records (EHRs) systems has increased tremendously, enabling the availability of patient health histories for analysis. EHRs are the digital version of a patient’s paper medical chart. The patient record contains detailed medical history, diagnoses, medications, treatment plans, laboratory test results, among other health information necessary for understanding a patient’s health status as well as their engagement with care providers within the inpatient, outpatient, and ambulatory care settings. Therefore, EHR data have been leveraged to address a variety of use cases including clinical research, quality improvement, clinical decision support, and population health. To support these use cases, computational methods are applied to lower-level tasks e.g., extracting medical events that can inform higher-level tasks, e.g., disease prediction (Solares et al., 2020). As of December 2019, there are slightly under 2.1 million papers published on EHRs for biomedical research (Shinozaki, 2019). By harnessing the power of EHRs, we could improve early-stage disease detection, risk prediction, and treatment evaluation; thus leading to a significant reduction in patient costs and improvement in patient outcomes.

The original purpose of recording medical data in EHRs systems; however, was primarily for billing and other purposes (Evans, 2016), not for research. Compared to using traditional paperwork, EHRs can help streamline healthcare delivery, thus improving quality of care (Menachemi and Collum, 2011). However, in the context of applying machine learning to solve problems for risk prediction, disease detection, and treatment evaluation, EHRs pose many challenges– they do not have a standardized format, can contain human errors and introduce collection biases. In addition, some institutions or geographic regions do not have access to the technology or financial resources necessary to implement EHRs, thus resulting in vulnerable and disadvantaged communities not being electronically visible (Shinozaki, 2019).

### 1.1 Defining Levels of Information Complexity for EHR Data

EHRs data used for research purposes typically require a great deal of preprocessing to generate a matrix form with well-defined features of a patient’s clinical case. To describe different levels of EHRs data, we suggest the following terminology: **Level 0** data refers to the raw data that resides in EHRs systems without any pre-processing steps. **Level 1** data refers to data that have been harmonized, curated, integrated, and standardized. **Level 2** data refers to EHRs data in matrix form that includes variables extracted through automated algorithms, chart reviews, etc (see Figure 1 for an example). Level 0 data lack structure or standardization (e.g., narrative text, non-codified fields). Level 1 data are complex – they oftentimes come in the form of a sequence of events with heterogeneous structure (e.g., templated text, codified prescriptions and diagnoses, lab tests, vitals, etc. to standard terminologies and vocabularies). In addition, data for individual patients are collected at irregular time intervals and frequencies. These challenges prevent researchers from being able to directly apply classical statistical and machine learning methods for analysis of Level 1 data. However, there is significant time spent pre-processing such as feature extraction/engineering through chart review or other means, and there may be substantial information loss to go from Level 1 to Level 2 EHRs data. Thus, for machine learning models to be adopted in a clinical setting, it is highly advantageous to build models that can use Level 1 data directly.

**Figure 1:**
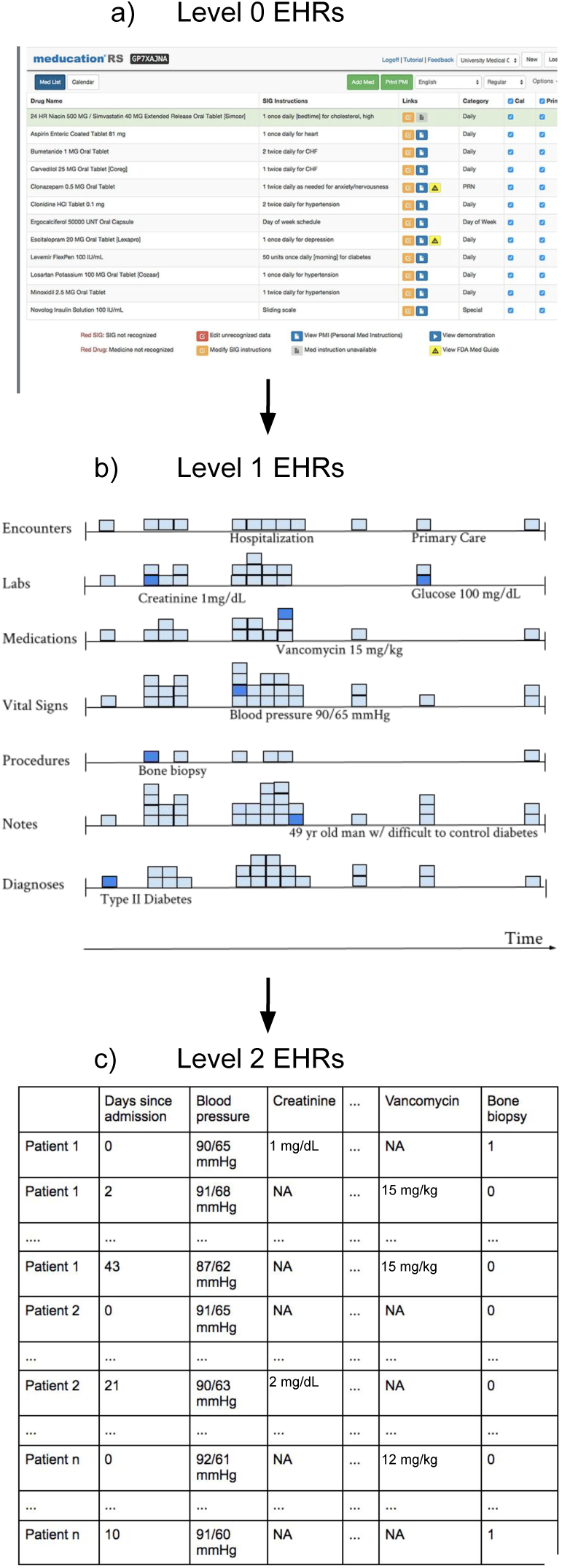
Different levels of EHRs data. a) displays Level 0 EHRs data (i.e., data still in EHR system without any pre-processing). b) displays Level 1 EHRs data after having been extracted from the EHR system and after minimal processing such as curation and harmonization. Note that Level 1 EHRs data are in sequential form with structured, codified, and unstructured data and no explicit missing values. c) displays Level 2 EHRs data that have been pre-processed into matrix form. Going from Level 1 EHRs data to Level 2 EHRs data requires significant effort spent pre-processing such as feature extraction/engineering through chart review or other means and may lead to substantial information loss. There are explicit missing values in Level 2 data due to its matrix form.

Individual patients typically have varying amounts of data in their EHRs for a variety of reasons – lack of collection / documentation, less medical visits, etc. (Wells et al., 2013). Many issues can contribute to biased data collection, particularly with regard to trends in health-decision making of different patients. For example, studies have found that individuals from vulnerable populations (e.g., immigrants, low socioeconomic status, psycho-social issues) are more likely to visit multiple healthcare institutions to receive care. In addition, low socioeconomic status patients may receive fewer diagnostic tests and medications for chronic disease (Gianfrancsco et al., 2018). Racial and ethnic minority groups have sub-optimal access to healthcare, with Black Americans in particular facing disparities in terms of health status, mortality, and morbidity (Hall et al., 2015). It has also been found that male patients tend to seek healthcare less than female patients (Loenen et al., 2015). Age also plays a role in health-seeking behaviors – studies found that adults 65 years or older tend to have more consultations with family physicians and undergo an annual health check (Deeks et al., 2009). The barriers that exist for young people seeking healthcare include discontinuities in care, lack of payment for transition support, and lack of preparedness for an adult model of care (Medicine and Council, 2015).

The consequence is that less data and information are documented in EHRs for patients belonging to certain groups due to fragmented care (Rajkomar et al., 2018). Models trained on data where certain groups are prone to have less data may exhibit unfair performance for these populations (Ghassemi et al., 2020). This can be viewed as a missing data problem. There is a growing recognition that ubiquitous missing data in EHRs, even when analyzed using powerful statistical and machine learning algorithms, can yield biased findings and unfair treatment decision, further exacerbating existing health disparities (Gianfrancsco et al., 2018, Rajkomar et al., 2018). Furthermore, when it comes to EHRs use for research, oftentimes investigators search for patients whose data are complete enough for analytical purposes (Weber et al., 2018), more likely excluding patients from under-served populations.

### 1.2 Assessing Impact of Missingness on EHR Data

Strategies for dealing with missing data tend to rely on assumptions about the nature of the mechanism that causes the missingness. As such, missing data are typically categorized into one of three classes: **Missing Completely at Random (MCAR), Missing at Random (MAR)**, and **Missing Not at Random (MNAR)**. MCAR, a term coined by Rubin, 1976, describes data where the probability of being missing is the same for all cases, thus implying that the cause of missingness is unrelated to the data itself. An example could be a lab technician forgetting to input data points into a patient’s health record, regardless of any attributes that describe a patient. The second class, Missing at Random (MAR), describes data in which missingness depends on the observed data. For example: a patient’s demographic characteristic is associated with seeking less healthcare and therefore having sparser medical records. MAR tends to be more general and realistic than MCAR. When data are MCAR or MAR, the response mechanism is termed ignorable, or a researcher can ignore the reasons for missingness in the analysis and thus simplify model-based methods (Heitjan and Basu, 1996). If neither MCAR nor MAR holds, the missingness is categorized as Missing Not at Random (MNAR). This means that the probability of being missing may depend on missing values (Buuren, 2018). This type of missingness can also be classified as non-ignorable if a patient’s unobserved underlying condition (e.g., undiagnosed depression) prevents them from traveling to see a healthcare provider (Schafer, 1997).

There is a growing body of work on handling missing data in health records, and as such there exists current methods for simulating missingness based on the three mechanisms. Beaulieu-Jones et al., 2018 outlined considerations for dealing with missingness in EHRs data based on their own numerical experiments. For their MAR simulations, missingness in a feature depends on another feature value. For MNAR simulations, missingness in a feature depends on its own value being in a selected quartile. Hubbard et al., 2018 conducted simulation studies to compare phenotyping methods to their Bayesian latent class approach. For MAR, missingness was simulated with a Bernoulli distribution based on patient-specific missingness probabilities varying based on age, race, and body mass index (BMI). For MNAR, missingness probabilities depended on Type 2 Diabetes Mellitus status. Both papers simulate missingness in Level 2 EHRs data, or EHRs data contained in a matrix with a priori defined features. It should be noted that from Level 1 EHRs data to Level 2, there is a sizable loss of information in addition to the labor-intensive process of manually and/or automatically converting to Level 2 data. Moreover, none of the existing works account for dependencies between EHRs events when artificially generating missing data in their experiments. An example of dependency is the relationship between diabetes and insulin. Most of the time, insulin is used to treat diabetes. Thus, if an instance of diabetes is removed, the insulin prescription should also be removed most of the time to reflect a realistic clinical scenario.

### 1.3 Modeling Medical Event Relatedness using Knowledge Graphs

To determine the relatedness between medical events, one might train a knowledge graph and then leverage its information to simulate realistic missingness. Knowledge graphs have previously been very useful in the analysis of electronic health records. Goodwin and Harabagiu (2013) automatically constructed a graph of clinically related concepts and presented an algorithm for determining similarity between medical concepts. Santos et al. (2022) developed a knowledge graph that integrates proteomics and clinical data in order to assist with clinical decision-making. Rotmensch et al. (2017) also developed knowledge graphs that link symptoms and diseases directly from electronic health records. They developed three types of graphs based on logistic regression, naïve Bayes classifier, and a Bayesian network using noisy OR gates. The constructed knowledge graphs were evaluated against Google’s manually-constructed knowledge graph and against physician opinions. They found that the noisy OR model produced a high quality graph that outperforms the other models. None of these methods have been applied for the purpose of accounting for event dependencies when simulating realistic missing data. We use the noisy OR-gate model from Rotmensch et al. (2017) in our experiments due to its high performance compared to naïve Bayes and logistic regression approaches, its flexible assumptions, and ease of use.

There is currently no framework for introducing realistic missing values in Level 1 EHRs data. There is also a gap in investigating the effects of missing data with regard to these data. As stated previously, Level 1 EHRs represent a sequence of events with heterogeneous structure with features that are not well-defined. This also includes repeated measures at different time points for different patients. Events are oftentimes coded multiple times in a single patient medical visit, and frequently there exists causal pathways between medical events in the patient’s record. As a result, we cannot rely on traditional methods to simulate realistic missing data in EHRs and assess their impact.

In this work, we develop an innovative framework to simulate missingness under the three mechanisms (namely, MCAR, MAR, and MNAR) that mimic real-life situations and health-seeking behaviors of various patients. We account for dependencies between medical events through the use of a medical knowledge graph. We create models for Level 1 EHRs for predicting future diagnoses at a patient’s next medical visit and quantify the extent to which prediction performance is affected by missing data under MCAR, MAR, and MNAR by the amount of missing data and by health-seeking behaviors that may lead to health disparities.

## 2 Materials and Methods

We choose to simulate missing data in the MIMIC-III (Medical Information Mart for Intensive Care III) dataset (Johnson et al., 2016). The MIMIC-III dataset consists of de-identified health-related data of patients who stayed in critical care units at the Beth Israel Deaconess Medical Center between 2001 and 2012. This dataset is a good Level 1 candidate for simulating missing data because the numbers of medical events are evenly distributed across most groups of interest. It is important to note that the MIMIC-III data is from the intensive care unit (ICU) and thus may be different from EHRs from outpatient and primary care providers. We are primarily interested in assessing the impact of missing data on patient groups that typically have less access to healthcare, and an evenly distributed dataset allows us to assess the impact by inducing more missingness in certain groups.

### 2.1 Simulating Realistic Missing Data within Patient Groups

We are interested in looking at four types of differences: gender, age, socioeconomic status, and race. We infer socioeconomic status based on insurance (government and private insurance). See Table 1 for average and median number of medical events per patient medical record across groups. We define younger patients (age 17-54), patients of color (Asian, Hispanic, Black, multi-racial, Middle Eastern, Hawaiian), male patients, and patients on government insurance as “data-lacking” groups as we would expect there to be more missing data in medical records from primary care providers and other types of EHRs due to either less access to care or seeking less care. We define older patients (age 55-70+), female patients, patients on private insurance, and White patients as “data-rich” groups due to such groups either having more access or seeking more care.

**Table 1:** Distributions of medical events across various demographic groups. Mean and median number of medical events per patient record are evenly distributed across age groups. There are more older patients than younger patients in the training data. We also see that mean and median number of medical events per patient record are evenly distributed across sex. Proportions of male-to-female patients are relatively even in the training data.

| Age | Mean Events | Median Events | Proportion of Patients |
| --- | --- | --- | --- |
| 17-55 | 333 | 254 | 0.257 |
| 55-70+ | 314 | 260 | 0.713 |
| Sex |  |  |  |
| Male | 321 | 261 | 0.537 |
| Female | 316 | 256 | 0.432 |
| Insurance Type |  |  |  |
| Government | 322 | 262 | 0.701 |
| Private or Self | 312 | 248 | 0.261 |
| Race |  |  |  |
| Black | 357 | 296 | 0.120 |
| White | 314 | 262 | 0.721 |

We extract the sequence of structured and codified events in the MIMIC-III database for each patient. Structured and codified events represented include prescriptions (p (drug names and NDC codes)), lab tests (l (identifiers associated with lab measurements; LOINC codes)), conditions (c ICD-9 codes), symptoms (s ICD-9 codes), and diagnoses (d ICD-9 codes). For example, d 250 represents diagnosis (d) of diabetes mellitus with an ICD-9 code of 250. Because we are evaluating a prediction tool to assist physicians with future diagnoses, we define the patient’s medical history in two ways: for patients with the diagnosis of interest in their records, we modeled the events in the medical visits prior to the current visit that contains the diagnosis of interest. For patients that do not have the diagnosis of interest, events in the visits prior to their most recent visit are used (see Figure 2). In this data structure, oftentimes structured and codified events are coded multiple times in the same visit.

**Figure 2:**
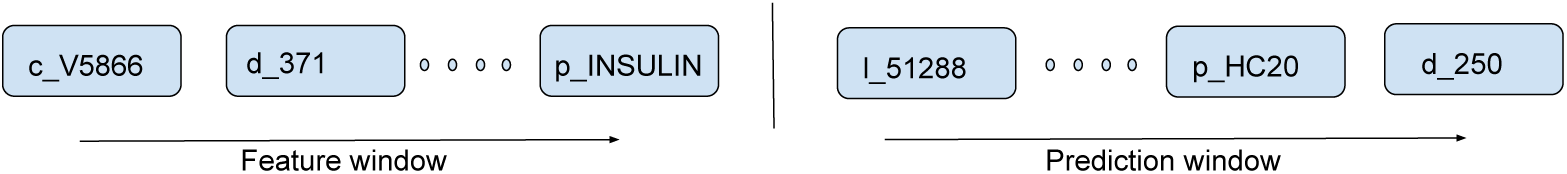
Sequence of structured and codified events used for prediction. Left panel refers to the events in the patient history, right panel refers to events in the next medical visit. Prefixes: c = condition, d = diagnosis, p = prescription, l = laboratory result.

### 2.2 Simulating Missing Data Under MCAR, MAR, MNAR

When data are Missing Completely at Random (MCAR), missing values are independent of both observed and unobserved data, and occur at random. In mathematical notation, we write this as:

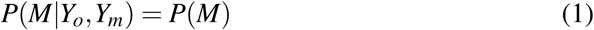

where *M* refers to the missing indicator, *Y*_*o*_ refers to the observed data, and *Y*_*m*_ refers to unobserved data. To induce realistic MCAR missingness, we mimic an EHRs situation in which a lab tech forgets to input data points. This means that even if a structured event is coded multiple times in the patient record, one instance of that event being removed does not affect the other instances. In our simulations, the number of patients that have clerical errors in their medical records (*X*_*i*_) follows a binomial distribution with probability 0.75 (*X*_*i*_ *∼ Bin*(*N*, 0.75)). For patients selected for clerical errors, we vary the proportion of events removed from each medical record from 0-0.75.

When data are Missing at Random (MAR), the events that lead to missingness are dependent on the observed data. In mathematical notation, we write this as:

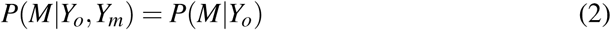

In our experiments, we explore variables such as sex, age, race, and insurance status (to infer socioeconomic status). We use a logistic regression model to simulate these conditional probabilities. In mathematical notation, this is:

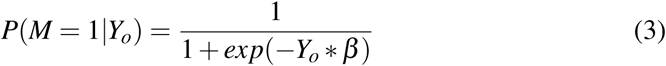

where *Y*_*o*_ now refers to our observed predictors (sex, age, insurance status, race) and *β* represents the coefficients for the predictors. We create models that reflect these differences in probabilities for these groups.

Since it has been shown that male patients are less likely to visit a healthcare provider than females, we select a *β* coefficient that reflect this difference in probabilities for sex. For insurance status, we separate patients on government insurance (Medicare, Medicaid) from patients with private insurance to represent lower and higher socioeconomic status. For race, we stratify Black and White patients. The *β*_*sex*_ coefficient is chosen such that *P*(*M*|*Male*) = 0.88 for male patients, and *P*(*M*|*Female*) = 0.35. We use the same *β* coefficient to determine probabilities for insurance status (with patients on government insurance having the higher probability for missing data) and race (with patients of color having the higher probability for missing data). For age, we treat this variable as continuous. We set our *β*_*age*_ such that the odds of having missing data are 12 percent higher for a decrease of one year in age. In Table 1, we display the different probabilities of being missing based on each covariate.

When neither MCAR nor MAR hold, the data are Missing Not at Random (MNAR). This means that after accounting for available observed information, the missingness still depends on unobserved data. In mathematical notation, this is:

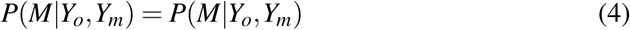

A realistic MNAR scenario in EHRs data is a patient’s unobserved underlying condition preventing them from going to health-related appointments (such as chronic pain, depression, etc). For each diagnosis, we try to predict in the next medical visit, we select three other conditions / diagnoses to be “underlying conditions”. To select these “underlying conditions”, we first isolate medical events that have a cosine similarity *>* 0.7, and manually investigate whether or not they could realistically be responsible for missing appointments. For example, if we are predicting a future diagnosis of Diabetes, we might find that ‘*d*_357’ (Inflammatory and toxic neuropathy) has a cosine similarity of 0.843 and ‘*c_V* 5867’ (Long-term insulin use) has a cosine similarity of 0.854. It is more likely that toxic nerve damage would prevent an individual from attending a medical visit due to the fact that it involves pain as opposed to long-term insulin use.

Table 2 displays diagnoses that we will predict in our numerical experiments, their accompanying “underlying conditions”, and corresponding cosine similarity.

**Table 2:** “Underlying conditions” for MNAR prediction models

| Target diagnosis | “Underlying condition” | Cosine similarity |
| --- | --- | --- |
| Diabetes | Toxic Nerve Damage | 0.843 |
|  | Fluid / Acid-Base Disorder | 0.817 |
|  | Anemia | 0.782 |
| Chronic Kidney Disease | Glomerulosclerosis | 0.877 |
|  | Nephritis | 0.855 |
|  | Impetigo | 0.771 |
| Heart Failure | Chronic Pulmonary Disease | 0.885 |
|  | Cardiomyopathy | 0.868 |
|  | Mitral Valve Disorder | 0.843 |
| Hypertensive Chronic Disorder | Toxic Nerve Damage | 0.720 |
|  | Fluid / Acid-Base Disorder | 0.720 |
|  | Chronic Pulmonary Disease | 0.719 |
| Lipid Metabolism Disorder | Hypothyroidism | 0.831 |
|  | Gout | 0.826 |
|  | Fluid / Acid-Base Disorder | 0.800 |

We use the same logistic regression model used in the MAR experiments with an added covariate for the “unobserved” underlying condition to determine probabilities for a patient to have missing data, or P(M | *Y*_*o*_, *D*_*i*_) where *D*_*i*_ is an indicator for having one of the specified underlying conditions. *D*_1_ corresponds to having at least one underlying condition in the medical history, and *D*_0_ corresponds to having no underlying conditions in the medical history. P(M | *D*_1_) = 0.88 and P(M| *D*_0_) = 0.35 in our model.

### 2.3 Applying a Knowledge Graph to Induce Realistic Missingness

To account for dependencies between EHRs events, we employ the use of a knowledge graph based on the noisy OR-gate model (Rotmensch et al., 2017). Noisy OR is a conditional probability distribution that describes causal mechanisms by which children nodes are affected by parent nodes. Parent nodes can be defined as diseases; children nodes can be defined as symptoms. In a deterministic noise-free setting, the presence of a disease would always cause symptoms to be observed, but in real life the process is less deterministic. The model deals with inherent noise in the process by introducing failure and leak probabilities. For example, the presence of a disease *y*_*j*_ might fail to turn on its child symptom *x*_*i*_ with probability 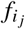. The leak probability *l*_*i*_ denotes the probability that a symptom is on even if all parent diseases are off.

The probability of a child being present is defined as

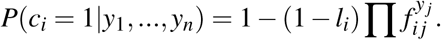

Then the importance measure is defined as

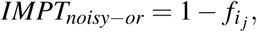

due to the fact that higher importance indicates that a disease is more likely to turn on or generate an observed corresponding symptom. Model parameters are learned using maximum likelihood estimation. The importance measure is derived from the conditional probability distributions, and thus no assumptions about the prior distribution of parent nodes are made. This model was shown in numerical experiments to outperform naïve Bayes and logistic regression models. Unlike other models, such as naïve Bayes and logistic regression, it does not make any assumptions about the prior distribution of outcomes. This is important since patients present with multiple types of medical events in their records.

In our experiments, we have five different event categories: diseases, abnormal lab tests, prescriptions, symptoms, and conditions. We determined that conditions would not be included in the knowledge graph due to the variety of non-clinical events that can be classified as a ‘condition’ in the MIMIC-III data set (for example, organ donor status). Thus, we learn four different noisy OR-gate models with each event category considered to be the parent node, and all other event categories considered to be the children nodes (see Figure 3).

**Figure 3:**
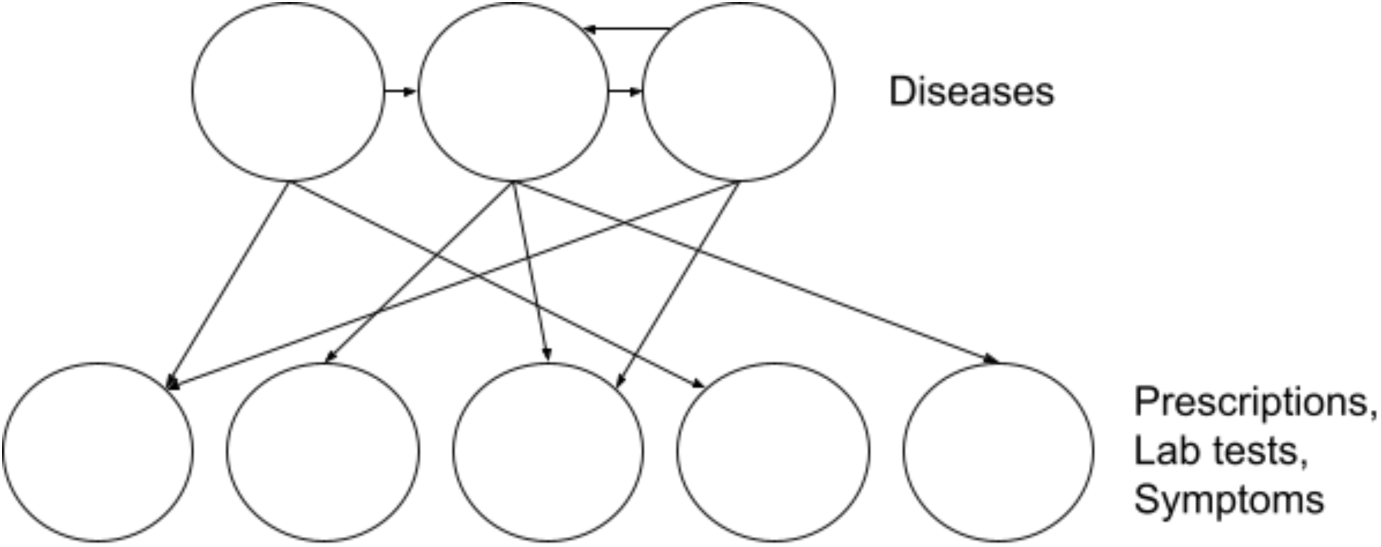
Noisy OR model example with diagnoses as the parent nodes and all other events as the children nodes

Because the MCAR experiments are based on clinical errors and do not capture any clinically meaningful situations, we do not use the knowledge graph for the MCAR event removal process. Instead, we simply remove events at random up to a certain threshold (which varies from 0-0.75 of events in a medical record if a patient is selected for missingness). See Figure 4 for the MCAR event removal process.

**Figure 4:**
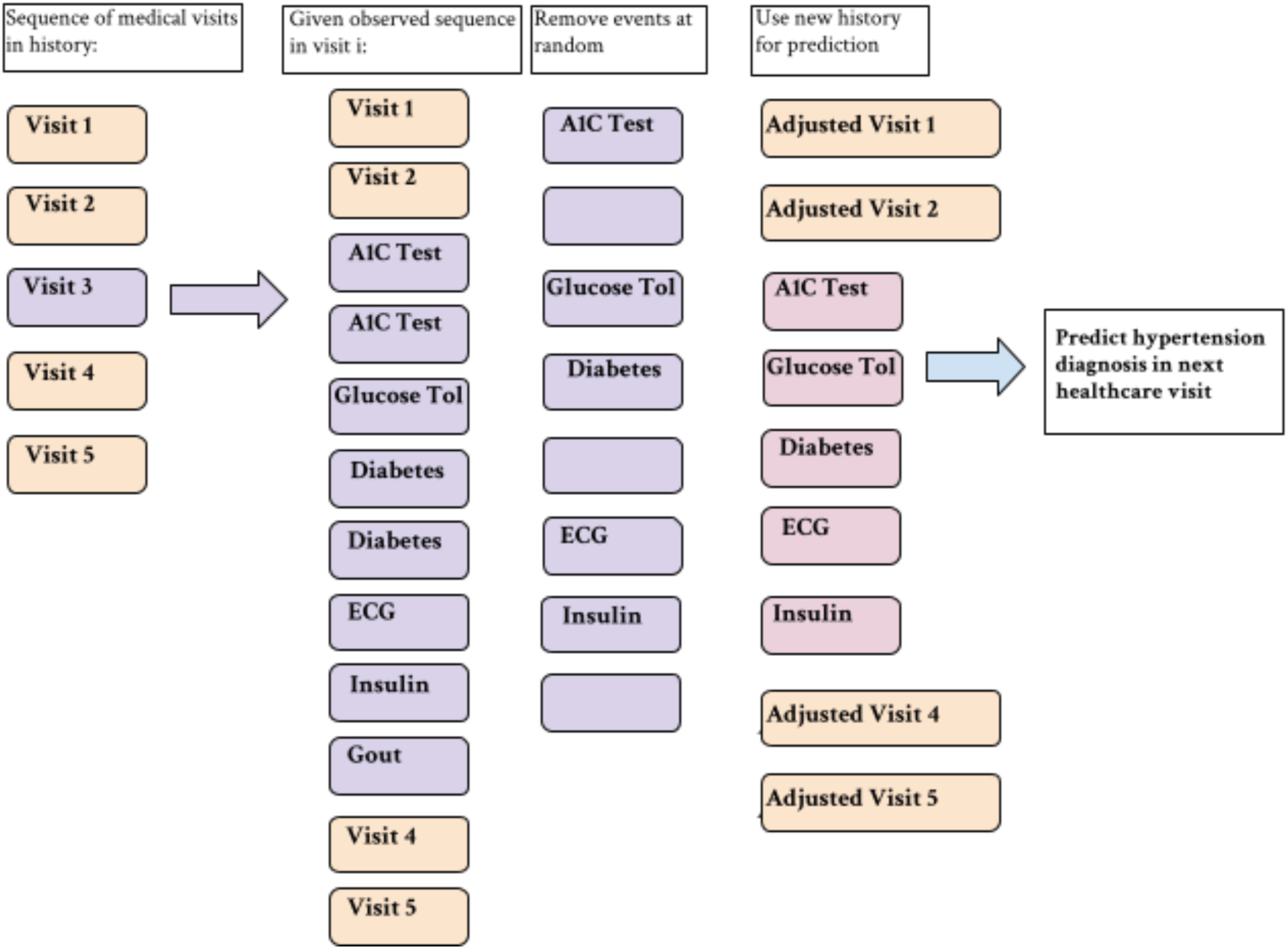
Clerical error simulation example. Events are removed at random, regardless of whether or not they appear multiple times in the record.

For the MAR experiments, a patient might miss a medical visit or have less diagnose/medication data due to having less access to or seeking less care. We first remove entire medical visits from the patient’s record, the proportion of which is varied from [0-0.48]. In the remaining visits, we remove individual medical events. In this type of situation, if a patient misses a lab test, the subsequent diagnosis and prescription that would have resulted from that abnormal lab test should also be removed. We utilize the knowledge graph to account for these dependencies.

Rather than determining a threshold on the importance measure to learn an edge for a knowledge graph, we take a more probabilistic approach for event-removal. The process for removing events is as follows: If event A is chosen for removal, we use the noisy OR-gate model that treats event A’s category as the parent node (for example, event A is a diabetes diagnosis, and thus we choose the model that treats diagnoses as the parent nodes, and treats prescriptions, lab tests, and symptoms as the children nodes). From there, each event in the patient’s record has a failure probability associated with it. The importance measure described above is calculated for each event (the likelihood that the diabetes diagnosis will “turn on” the event). We treat each corresponding importance measure as the probability that the corresponding event will also be removed with event A. Of the events removed using this method, the event with the highest importance measure is chosen as the new parent node and its corresponding importance measures for the remaining medical events are calculated and the process repeats. If no additional events are removed, a random medical event is chosen and the process repeats. This occurs until reaching a certain threshold of events removed. At the highest threshold, the proportion of data removed from the medical record averages at *∼*0.75. See Figure 5 for the MAR event removal process using the medical knowledge graph.

**Figure 5:**
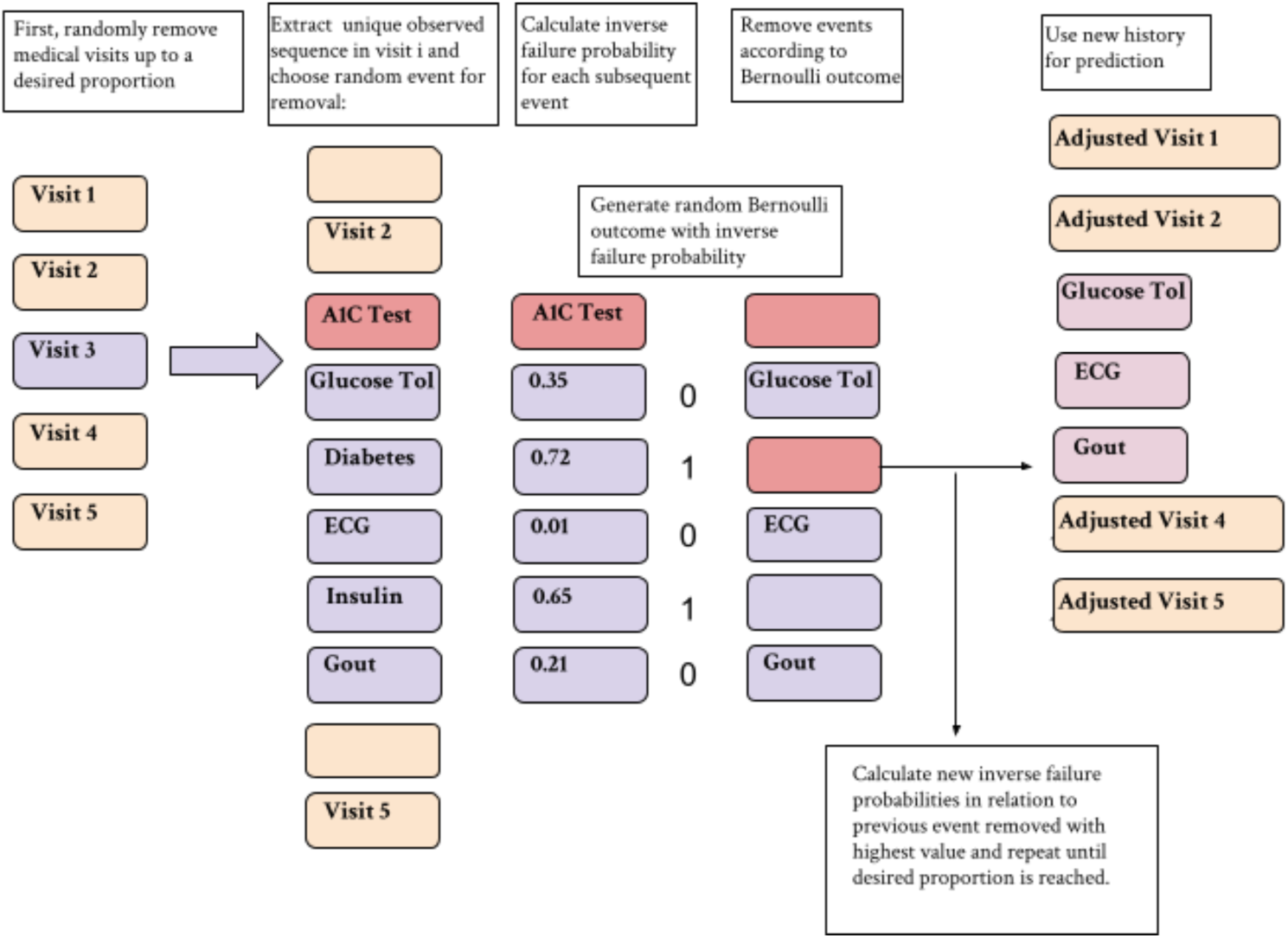
Event removal process for MAR experiments that incorporate the use of the noisy OR-gate knowledge graph.

We also use the knowledge graph for removing events in the MNAR experiments.

## 3 Numerical Experiments

In our numerical experiments, we focus on assessing the impact of missing data in EHRs on prediction accuracy. To build disease prediction models using EHR data, we applied a two-step approach.

### 3.1 Modeling Disease Prediction based upon Medical Histories

In the first step, we generate an embedding for the medical history of each patient by leveraging word embedding algorithms (Farhan et al., 2016; Getzen et al., 2022)). Word embedding algorithms can be extremely useful in the analysis of Level 1 EHR data. These algorithms convert words to vectors of real numbers such that the cosine distance between two vectors reflects the similarity of two words. In using the structured and codified data from Level 1 EHR data for word embedding, one can think of a “word” as a medical event in the patient record, and a “sentence” as the chronological sequence of all events in a patient record. We use the algorithm Word2Vec (Mikolov et al., 2013) to generate embeddings for medical events. Word2Vec has two training mechanisms: continuous bag of words (CBOW) and skip-gram. CBOW utilizes the surrounding context (other words around the target word) to predict a target word, and skip-gram uses a single word to predict a target context. The hidden layer following training contains the vectors that correspond to each word (Figure 6).

**Figure 6:**
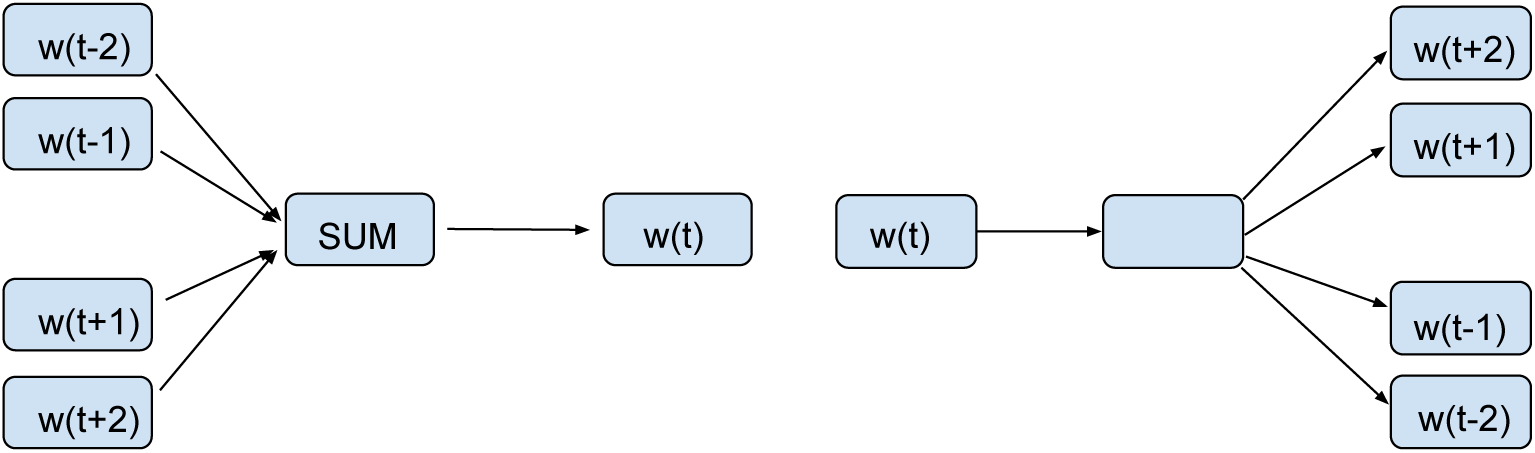
Training mechanisms for FastText – CBOW (left) and Skip-Gram (right). CBOW trains by using a context to accurately predict a target word, Skip-Gram trains by using each word to predict its target context.

### 3.2 Developing and Evaluating Disease Prediction Models

In the second step, we build three models for predicting a diagnosis of interest in a future medical visit, including diabetes (prevalence = 0.318), chronic kidney disease (prevalence = 0.203), heart failure (prevalence = 0.371), hypertensive chronic disorder (prevalence = 0.180), and lipid metabolism disorder (prevalence = 0.275). The ICD-9 codes are generalized to level 3.

Once we generate embeddings for individual events in a patient’s medical history, we derive an embedding for the patient’s entire medical history by multiplying each individual embedding by a temporal factor and summing up the vectors in the patient record. The equation that represents this process is as follows:

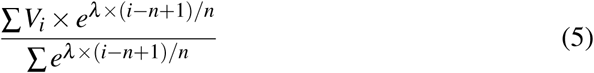

where *V*_*i*_ is the embedding of event *i* in a patient’s medical record, *λ* is the time decay factor, and *n* is the total number of events in the record. For our experiments, a decay factor of 5 is used. Figure 7 is useful for visualizing this process.

**Figure 7:**
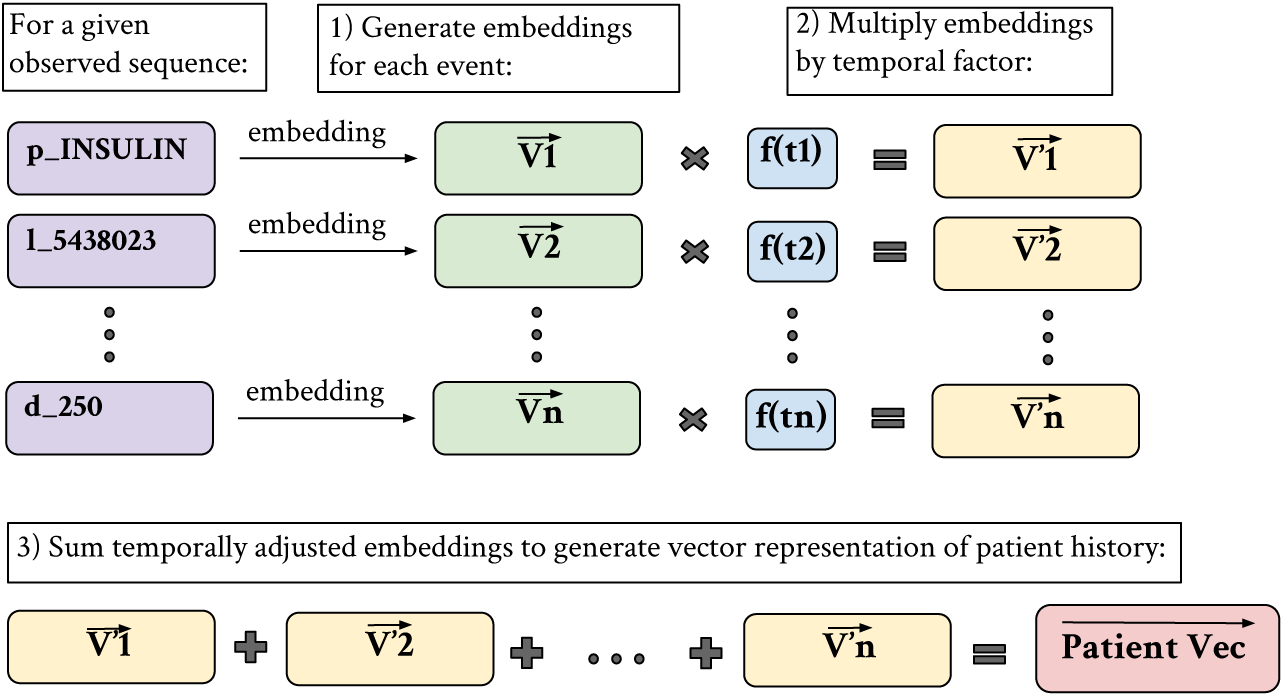
Patient vector generation process– embeddings are generated for each medical event in a sequence. Each embedding is multiplied by a function of its ordering, such that more recent events have more importance, and the temporally adjusted embeddings are summed to create an overall patient vector.

We develop and evaluate prediction models using three approaches: patient diagnosis projection similarity (PDPS), lasso (least absolute shrinkage and selection operator) regression, and artificial neural network.

Patient Diagnosis Projection Similarity (Farhan et al., 2016) involves projecting patient sequences into the vector space while accounting for the temporal impact of events, as described previously. The cosine similarity between the patient vector and the diagnosis of interest is used to predict whether or not a patient is at risk for developing that disease. One of the main benefits of the method is its ability to predict risks for multiple diseases simultaneously. We apply PDPS to evaluate the effects of missingness on results (see Figure 8).

**Figure 8:**
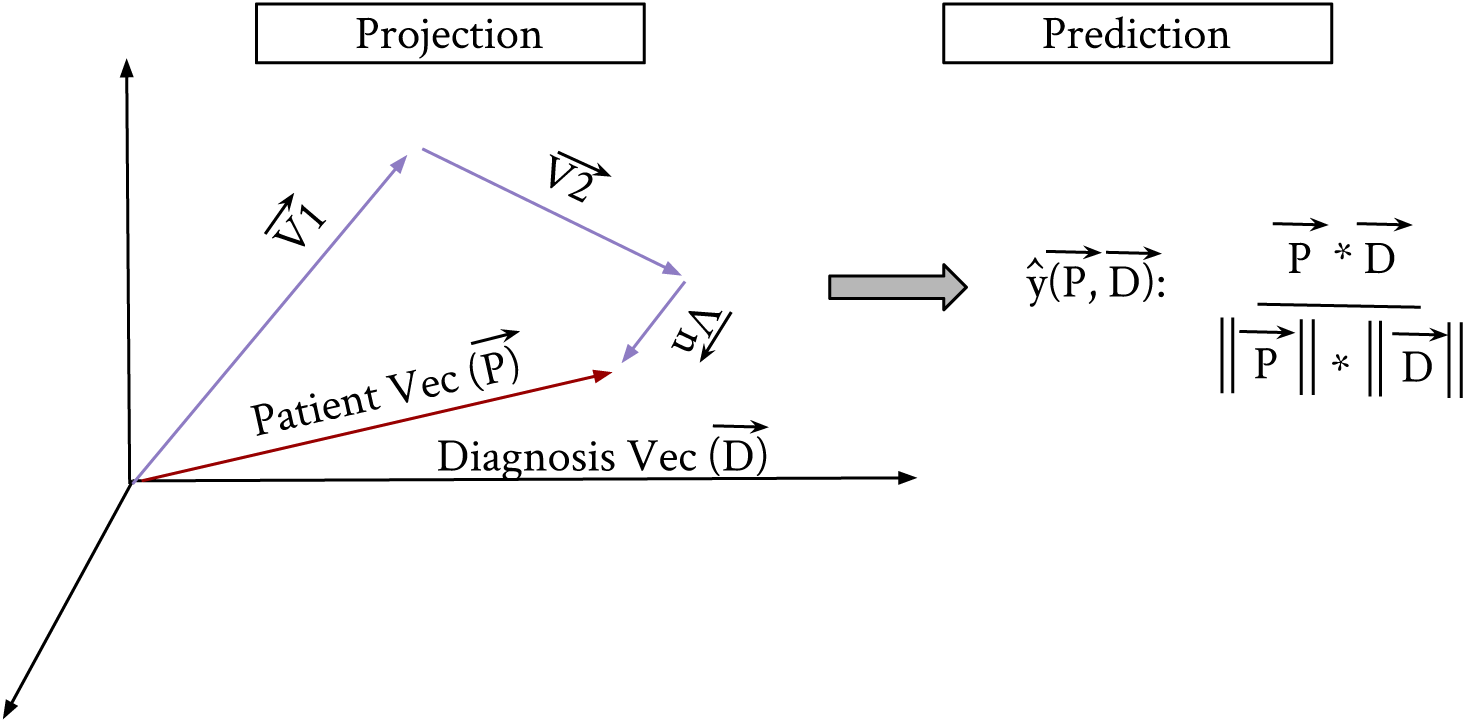
Patient Diagnosis Projection Similarity by Farhan et al., 2016. Patient vectors and target diagnosis vectors are projected into the vector space. Prediction is based on cosine similarity between vectors.

As an alternative approach, we also leverage the patient vectors to build disease-specific prediction models by using lasso (least absolute shrinkage and selection operator) regression and deep learning based on a three-layer artificial neural network with mean-squared error loss. Each coordinate in a patient vector can be viewed as a feature used for the prediction of a medical outcome. Figure 9 displays an example of this data structure: each row in the matrix corresponds to a patient vector, and each column corresponds to a coordinate feature. The last column indicates whether or not the patient is labeled as positive or negative for having a diagnosis of interest in a future medical visit.

**Figure 9:**
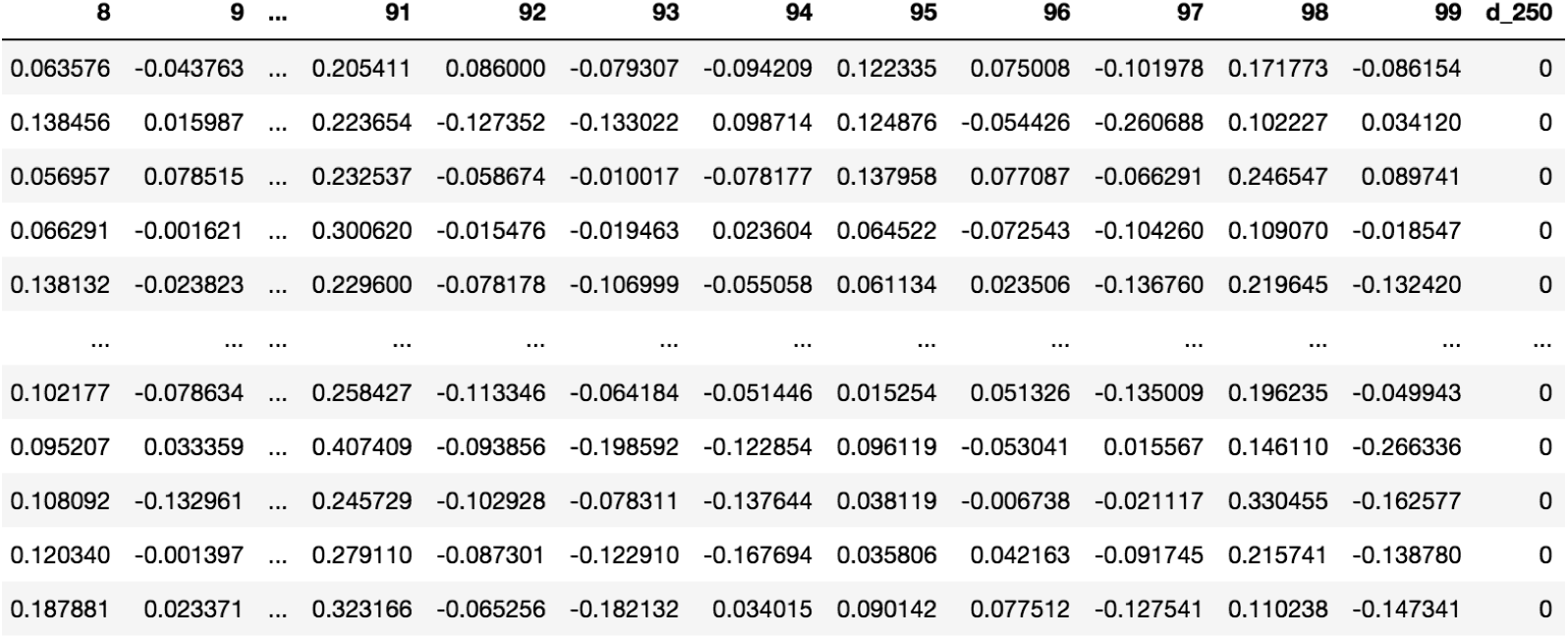
Sample patient vector data matrix. Each row in the matrix corresponds to a 100-dimensional patient vector. Each coordinate can be used as a feature for prediction of a future diagnosis (Diabetes in this example).

We evaluate model performance by area under the operating curve (AUC) at various missingness proportions for the patients selected to have missing data by their given model. There are two scenarios for this. In the first scenario, we only remove medical events from the training data and leave the testing data complete. This is a useful approach because it provides a consistent benchmark to compare results at different missingness proportions; however, it does not reflect a realistic situation in which the training and testing data come from similar populations. In the second scenario, we remove medical events from patients in both the training and testing data. Evaluating with different testing sets means that the subsequent results are not directly comparable; however, the changes in prediction performance would reflect more of a real-life situation where models are trained and evaluated with missingness incorporated.

In each experiment, we report the results that are averaged over 200 Monte Carlo datasets. We present our experiments evaluated with the neural network in the main results (Figures 10-17). Since the results for lasso regression were nearly identical, we do not include them. PDPS performed very poorly (AUC averaging at 0.5 for all diagnoses), possibly due to the fact that in this work we remove visits that contain the target diagnosis in the patient history. For this reason, we do not present these results.

**Figure 10:**
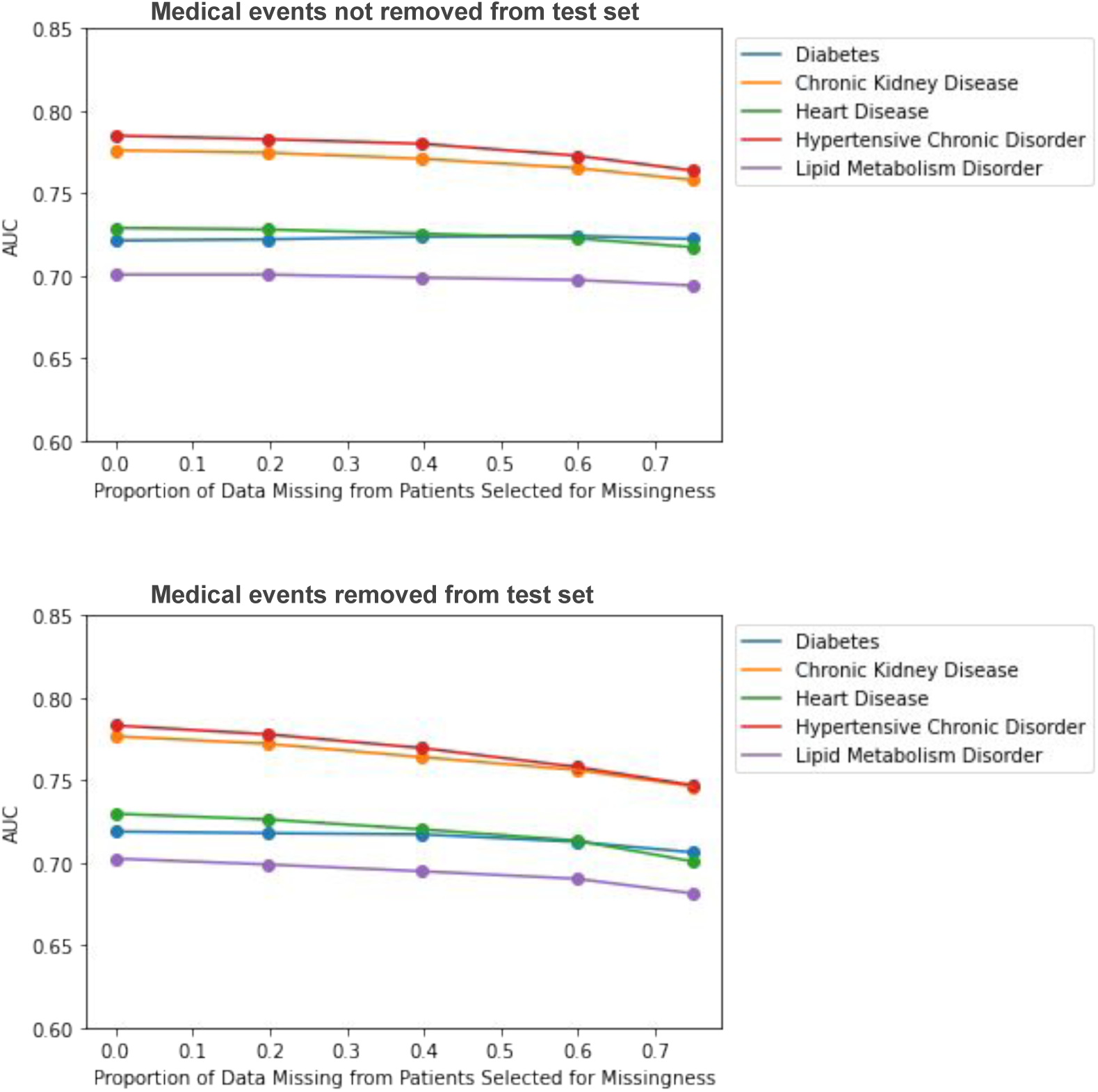
Impact of MCAR missing data on disease prediction model performance. Top figure corresponds to complete test set experiments, bottom figure corresponds to incomplete test set experiments.

## 4 Results

We outline the key findings of disease prediction accounting for three types of missingness: MCAR, MAR, and MNAR among data-lacking and data-rich patient groups.

### 4.1 Assessing Data Missingness across Disease Types

In the first set of results, we look at five disease prediction models (diabetes, chronic kidney disorder, heart disease, hypertensive chronic disorder, lipid metabolism disorder). The models are evaluated at increasing proportions of missingness in patients selected to have missing data in their medical records. We look at two scenarios: medical events removed only from the training set, and medical events removed from both the training and testing data. The former has the advantages of being able to evaluate on the same test set each time; however, the latter reflects a more realistic scenario in terms of evaluating the impact of missing data on underserved populations.

In the MCAR experiments, we randomly choose 78 percent of patients to have missing data to determine the extent of the impact of missing data on prediction results. If a patient is chosen to have missing data, we remove events at random from their medical history. The model performances when missingness is only induced in the training data does not diminish very much. When missingness is simulated in both the training and testing data, we observe a slight gradual decline in AUC for predicting all diagnoses at increasing levels of missingness (Figure 10).

In the MAR experiments, we use a logistic regression model with four covariates (age, sex, insurance status, race) to determine whether or not a patient is selected to have missingness simulated in their medical record. ∼ 78 percent of patients overall were selected for missingness based on this model. Missingness is induced by removing visits and unique events at random for selected patients. If a patient is selected to have a visit removed, and they only have one visit in their history, they are removed from the analysis. However, their data is factored into the proportion of missing data. We first assess the impacts of missingness on all patients in our data. We see a stronger decline in performance as the proportion of missing data increases when we remove events from both the training data and test data as opposed to just the training data (see Figure 11). We also see a slightly stronger decline in performance as we increase the proportion of missing data for MAR experiments compared to MCAR.

**Figure 11:**
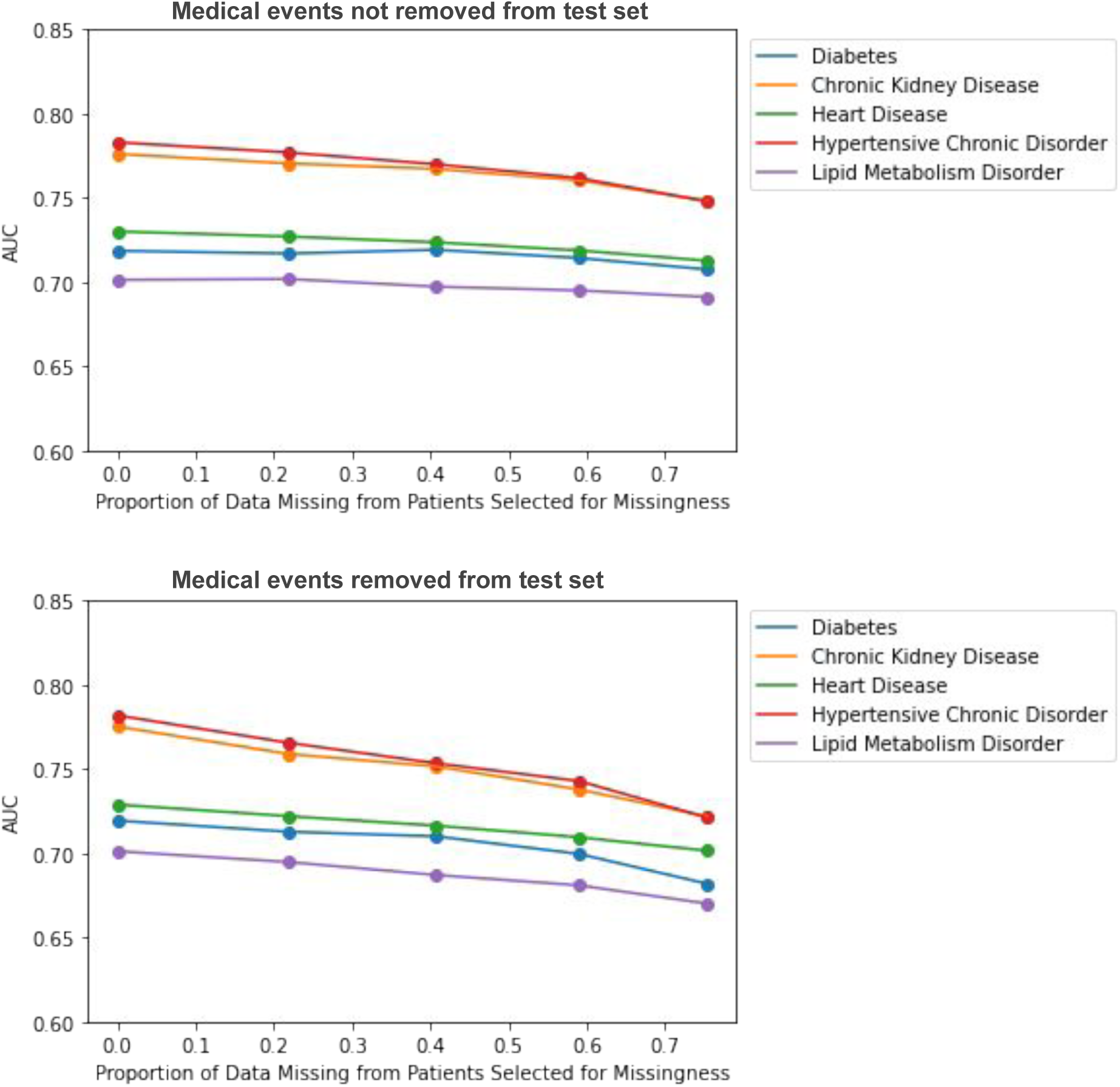
Impact of MAR missing data on disease prediction model performance. Top figure corresponds to complete test set experiments, bottom figure corresponds to incomplete test set experiments.

In the MNAR experiments, we use the same model as MAR, but include an additional covariate that accounts for an “unobserved underlying condition”. We choose a *β* coefficient such that P(M) = 0.88 for patients with a selected underlying condition, and P(M) = 0.35 for patients without. If a patient is selected for having missing data and the conditions are present in the record, the conditions are also subsequently removed after missing data is simulated. The overall effect looks similar to the MAR experiment, with declines in model performance as proportion missing in those selected for missingness increases (see Figure 12) The declines are stronger than in the MCAR experiments.

**Figure 12:**
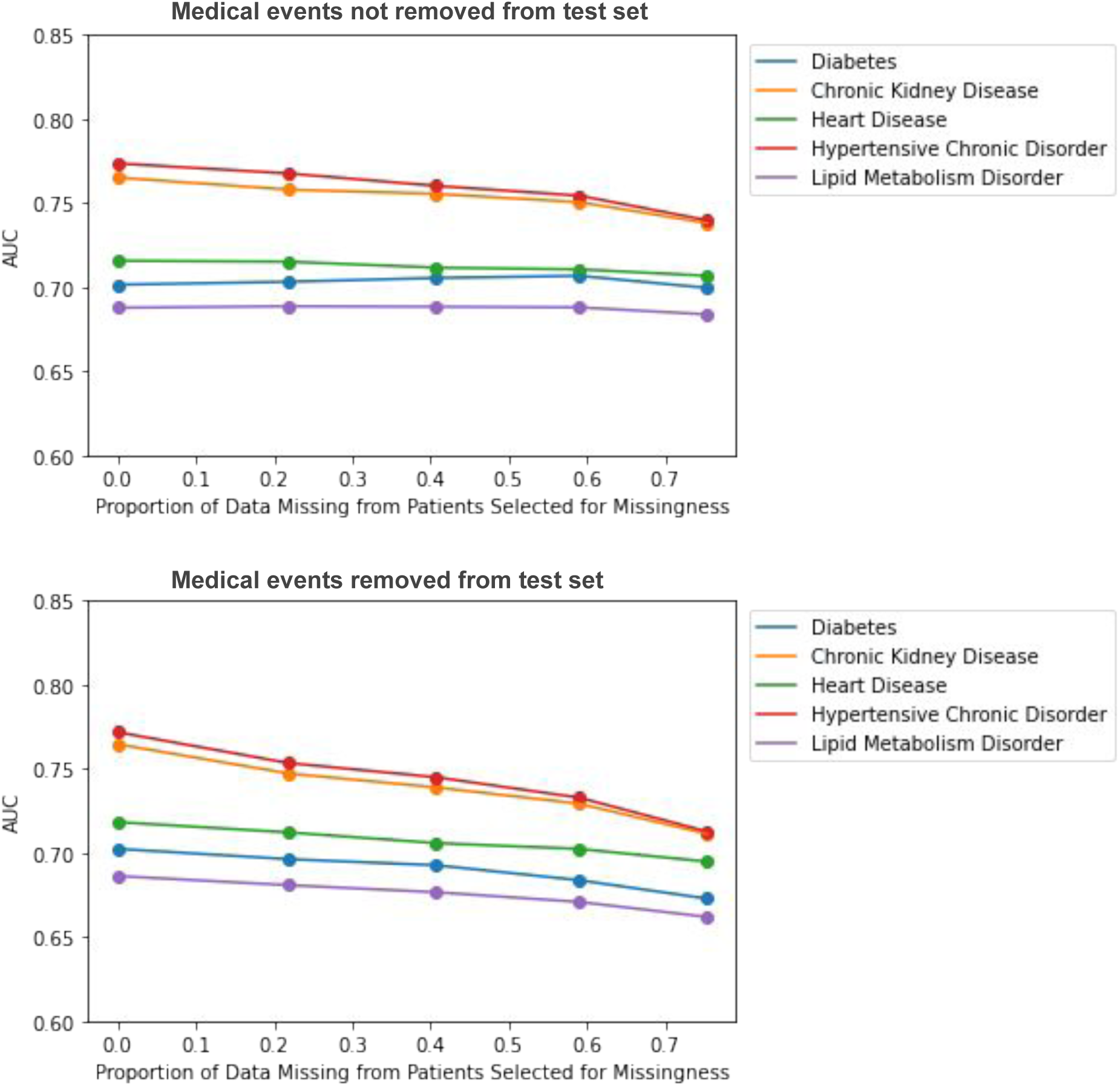
Impact of MNAR missing data on disease prediction model performance. Top figure corresponds to complete test set experiments, bottom figure corresponds to incomplete test set experiments.

For the complete test set experiments, we see that model performance remains more constant across different levels of missingness if missingness is not simulated in the test set, compared to when it is. A possible reason for this could be the fact that multiple similar medical events are used to predict a future diagnosis, and these events would always be present in the test set. For the incomplete test set experiments, we see stronger decreases in model performance for each missingness mechanism.

In the incomplete test data MCAR experiments, we do not observe as notable of a decrease as we do for MAR and MNAR. This is expected because events can be coded multiple times in an EHRs record, so to remove an event at random does not necessarily mean it does not exist elsewhere in a patient’s record. Dependencies are also not removed and thus likely help improve prediction performance since similar events probably have similar vector representations. In the MNAR models, the model performances across various levels of missingness look very similar to the MAR model performances. Thus, it appears that the addition of an underlying condition covariate does not contribute to further declines in disease prediction performance. This is surprising due to the fact that the underlying conditions were chosen such that they would be associated with the diagnosis of interest that we were trying to predict.

### 4.2 Assessing the Impact of Data Missingness for Data-Lacking Groups

Because we are interested in assessing the impact of missingness on populations with less access to healthcare or behaviors leading to less engagement with the healthcare system (data-lacking groups), in the next set of numerical experiments for MAR, we stratify by demographic group by splitting the testing data to evaluate each group separately. All patients begin with an average of 15 percent of their data missing. Patients that belong to a particular data-lacking group will have this average increase in increments. Patients that belong to the data-rich group will have this average stay at 15 percent. The idea is that we want to determine how more missing data in data-lacking groups affects their model performance, and compare it to groups that are not as affected. We do four different versions of this to look at the effects of insurance status, age, and race independently. In these experiments, we only present experiments where the events are removed from both the training and the test set (see Figures 13-15).

**Figure 13:**
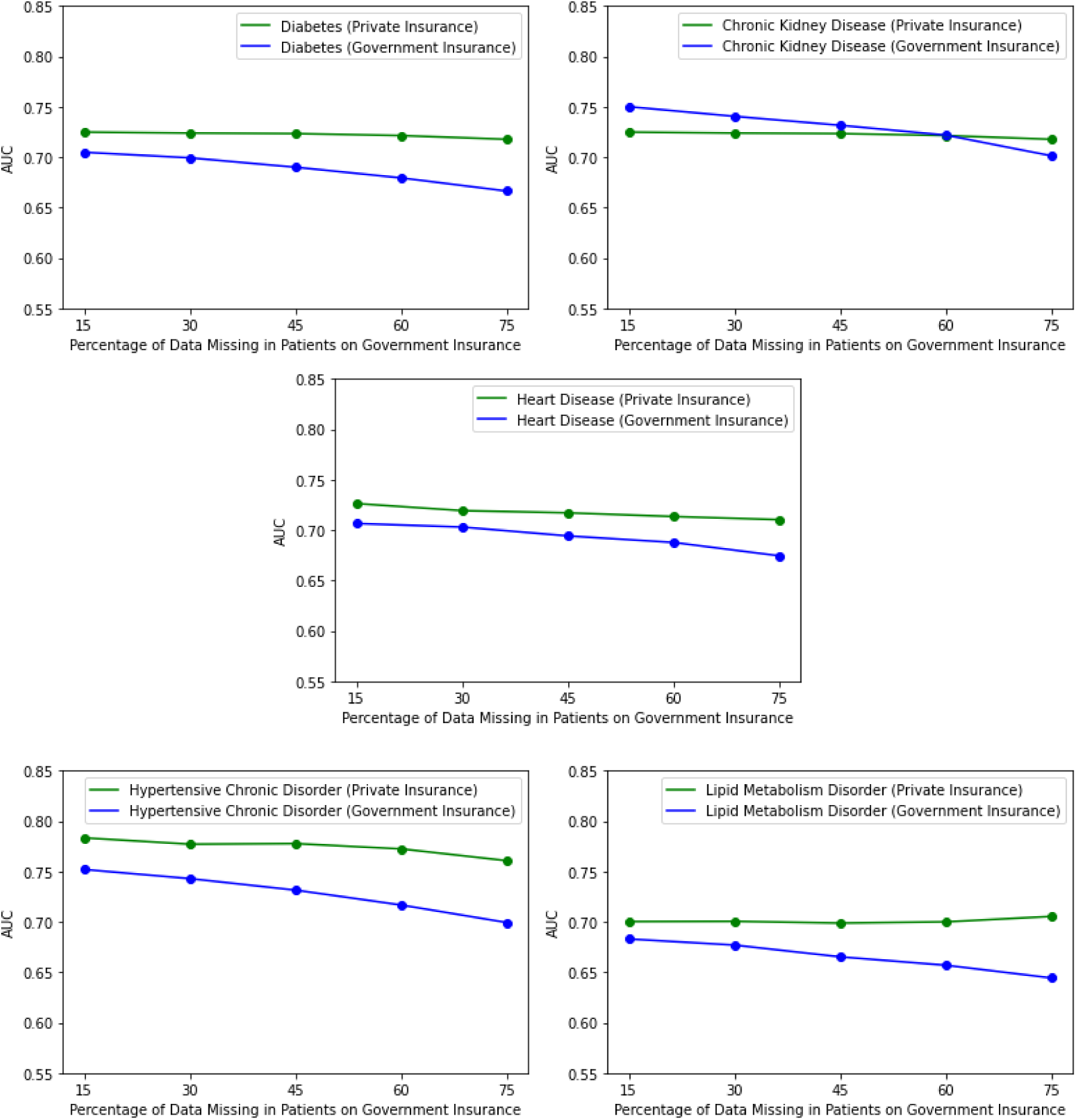
Assessing the impact of missing data on data-lacking groups. Proportion missing for patients on private insurance is held constant while proportion missing for patients on government insurance is varied.

First, we stratify our test set by insurance status. In the logistic regression model, we chose a *β* coefficient such that P(M) = 0.35 for patients on private / self insurance and P(M) = 0.88 for patients on government insurance to reflect less access to healthcare. We see initial disparities for patients on government insurance in predicting all diagnoses except for chronic kidney disease. As proportion of data missing increases for patients on government insurance, we see a stronger decline in model performance at the various increments. We also stratify our test data by younger patients (*<* 55) and older patients (*>* 55). We see that initially the AUC is higher for predicting all diagnoses for younger patients as opposed to older patients (Figure 13). As missingness in younger patients increases, we see declines in model performance, but it only leads to worse performance in predicting lipid metabolism disorder at the highest proportion of missing data (Figure 14). Lastly, we stratify by White patients vs. Black patients to see how race plays a factor in impact of missing data. We see strong initial disparities in predicting hypertensive chronic disorder for Black patients. We see very strong declines in model performance for predicting Diabetes as the proportion of missing data increases. We do not see these declines when predicting heart disease and lipid metabolism disorder (Figure 15).

**Figure 14:**
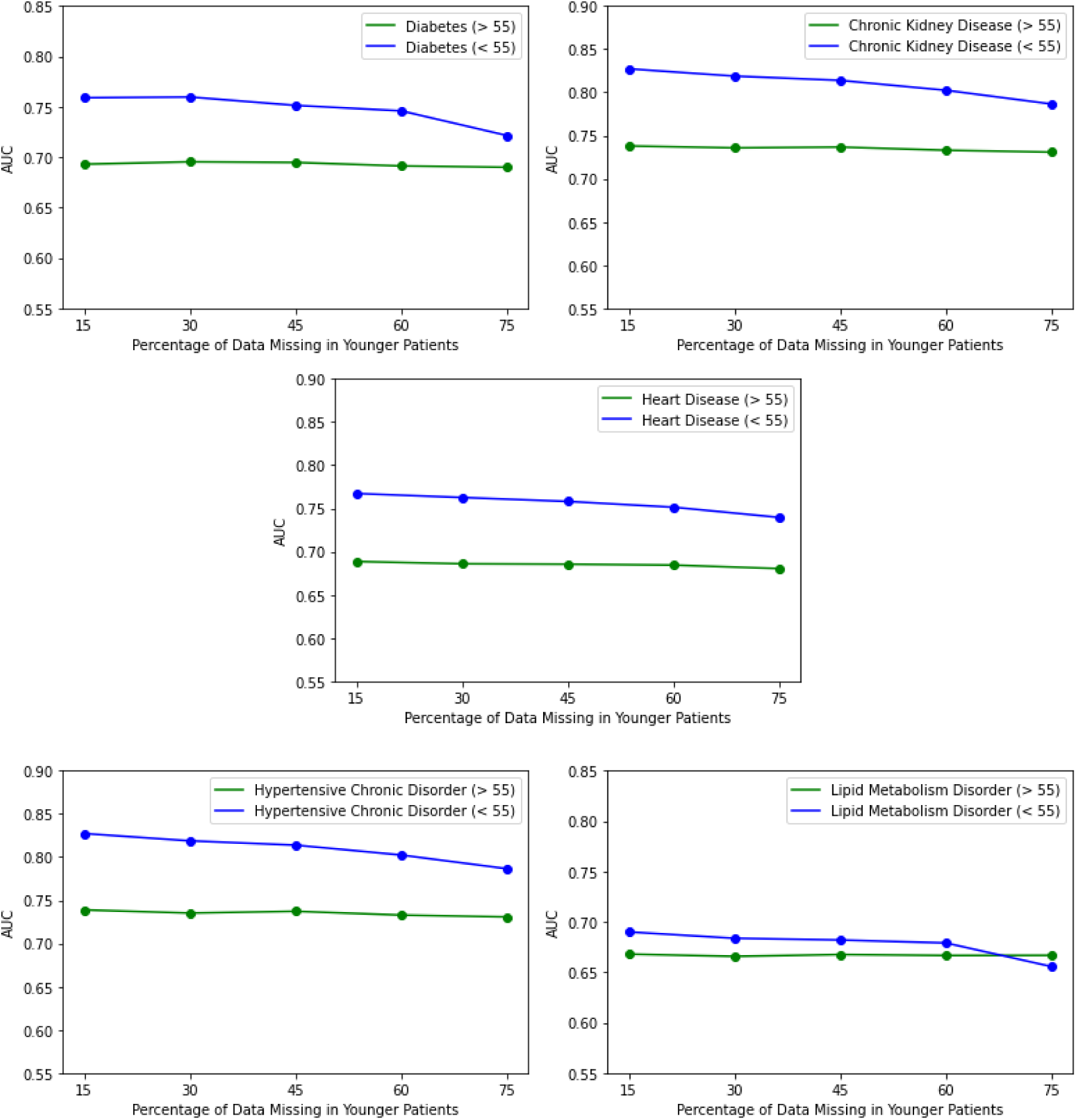
Assessing the impact of missing data on data-lacking groups. Proportion missing for older patients is held constant while proportion missing for younger patients is varied.

**Figure 15:**
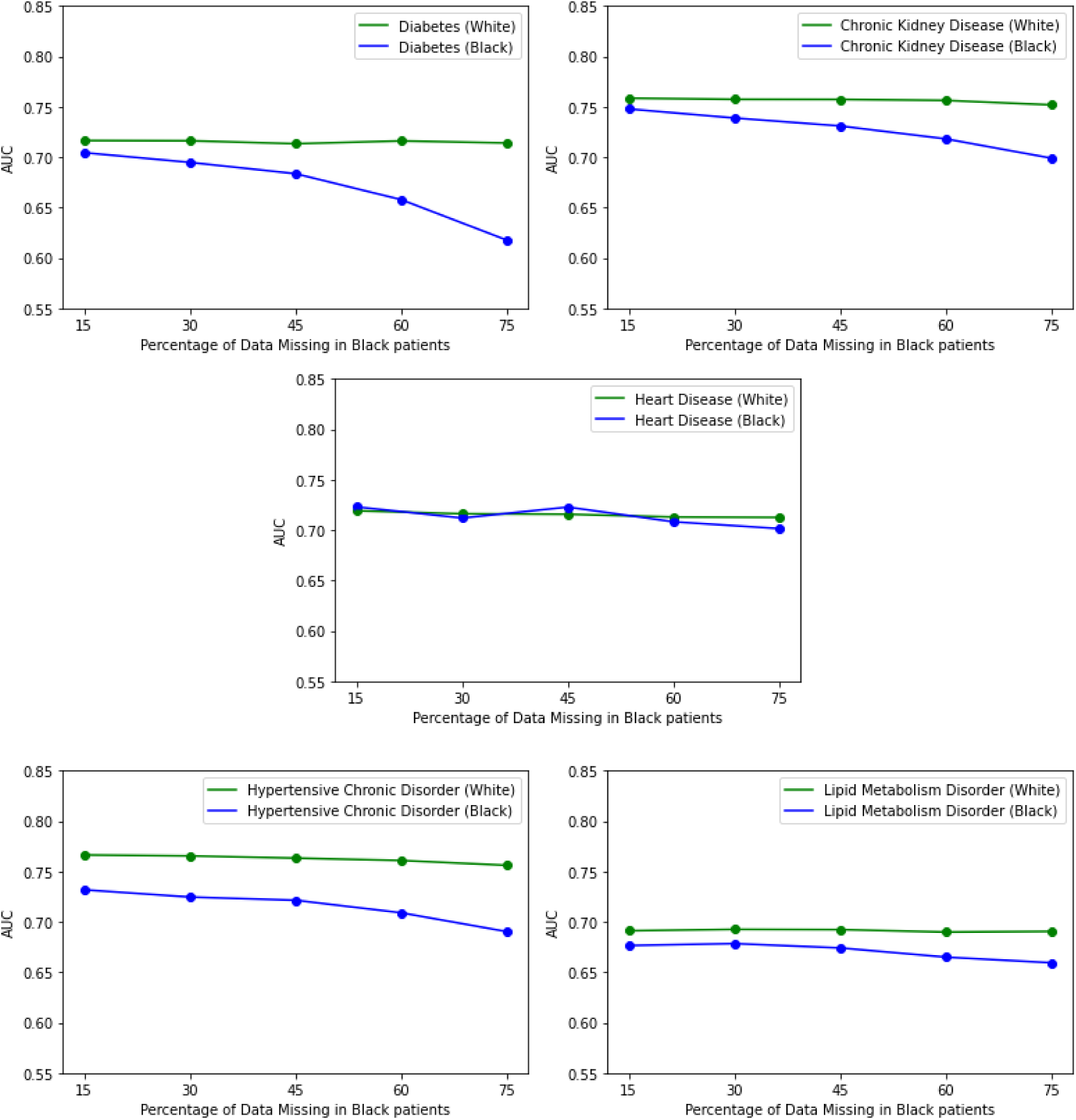
Assessing the impact of missing data on data-lacking groups. Proportion missing for White patients is held constant while proportion missing for Black patients is varied.

When assessing the impact of missing data on data-lacking groups, we observe that for most diagnoses, model performance for such groups are negatively impacted by missing data. Notably, some model performances for certain diagnoses are more affected by missing data than others. Models for diabetes, chronic kidney disease, and hypertensive chronic disorder are all more affected by missingness than models for heart disease. One possible explanation for this phenomenon could be that there are fewer types of events at smaller frequencies that are predictive of diagnoses like diabetes, chronic kidney disorder, and hypertensive chronic disorder. On the other hand, there could be many events at high frequencies that are predictive of heart disease, thus causing the model to not be as affected by missing data. We also see that certain data-lacking groups are more affected by missing data than others (for example, the decline in performance for younger patients as missingness increases is not as strong as the decline in performance for Black patients for certain diagnoses). Due to differences in quality of care, it is possible that Black patients have less predictive events for these diagnoses as well.

### 4.3 Comparing Event Removal with and without a Knowledge Graph

Next, we directly compare scenarios where all patients are selected for missingness and either have events removed at random (like in the MCAR experiments) or events removed using the knowledge graph (removing clusters of related events together) (Figures 16-17). In these experiments, all patients have missing data. We vary the proportion of the amount of missing data in each patients’ record to assess the impact as we did before. In these experiments, entire visits are not removed as they are in the MAR experiments. We simply use the knowledge graph event-removal methodology from section 2.3 to remove random clusters of dependent events from each medical record.

**Figure 16:**
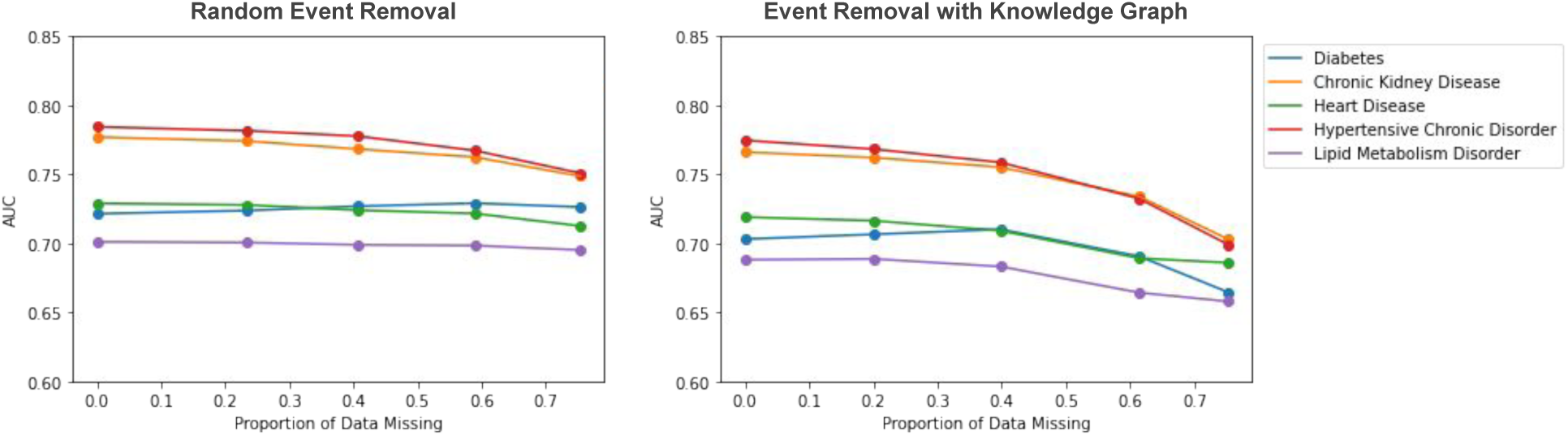
Impact of knowledge graph in event removal. Events are only removed from the training data. On the left, we assess the impact of removing medical events at random. On the right, we assess the impact of removing medical events using the knowledge graph.

**Figure 17:**
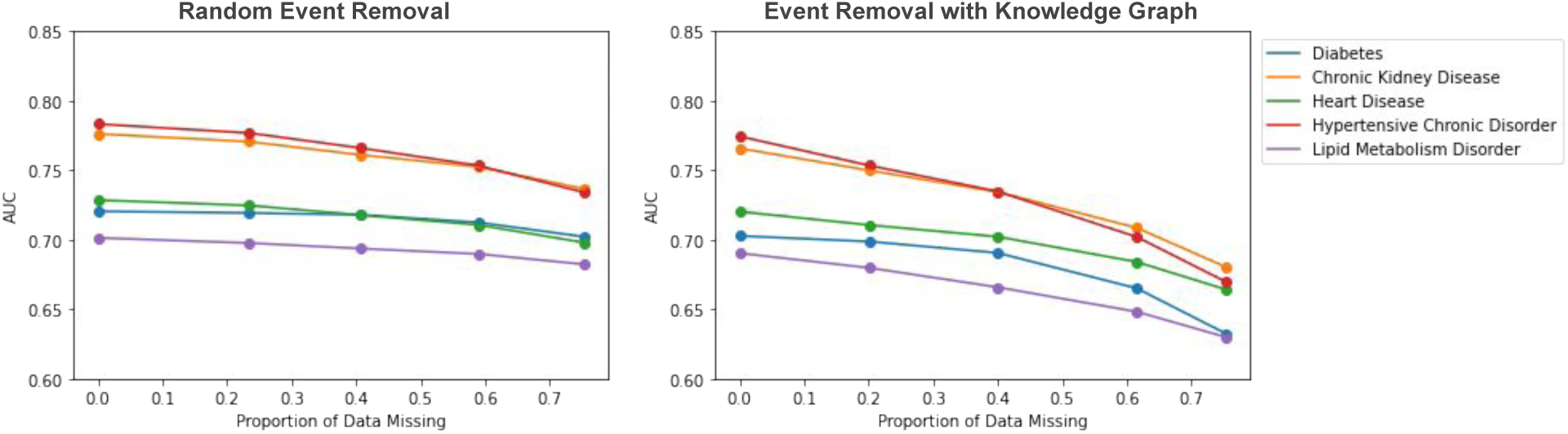
Impact of knowledge graph in event removal. Events are removed from both the training data and testing data. On the left, we assess the impact of removing medical events at random. On the right, we assess the impact of removing medical events using the knowledge graph.

We observe that in both scenarios (removing events from only the training data and removing events from all data), there are notably stronger declines in model performance as the proportion of data missing increases if events are removed using the medical knowledge graph.

## 5 Discussion

In this work, we propose a novel approach for simulating realistic missing data in Level 1 EHRs and evaluate the impact of such missing data on downstream leaning tasks through the use of prediction models that use the entire patient history. This general approach can be adapted and refined. For example, we used the noisy OR-gate model to obtain a knowledge graph for medical events in our numerical experiments. Alternatively, we can use knowledge graphs from other existing databases for medical events to capture dependencies and pathways for simulating missing data or other research purposes. In our work, we observe diminished model performance as a result of all types of missingness, and this diminished performance is more subtle in some scenarios (MCAR) as opposed to others (MAR, MNAR). We also see that disease prediction performance for data-lacking groups is more significantly impacted by missing data. Lastly, we find that for the same amount of missing data, accounting for relatedness of medical events and concepts in EHRs leads to greater impact on model performance.

These results indicate that disproportionate missing data in patients in certain demographic groups does impact disease model performance in a negative way. What this means is that the same disease prediction model may not be as effective for patients that have less data in their medical records. This has real implications for patients in under-served populations, particularly patients of lower socioeconomic status and patients of color – as advances in AI are incorporated into the clinic, these groups could get left behind from progress in early disease detection and risk prediction that these technologies afford. The divide in health outcomes with regard to attributes such as socioeconomic status and race is already wide. If we were to naively implement and integrate these models into the clinic, the gap could widen even more. Thus, it is of utmost importance to address missing data in EHRs for under-served populations when applying powerful ML methods. As scientists and engineers, it is our responsibility to ensure that our contributions do not leave people behind.

Dependencies exist between EHRs events in the real world. Diagnoses typically happen as a result of abnormal lab tests, and prescriptions typically occur to treat a diagnosis. Although we cannot know the exact causal pathways between these events, we can do a better job in accounting for these correlations when simulating missing data in Level 1 EHRs. The difference in results when we simulate missing data via knowledge graph compared to random event removal displays the importance of generating realistic missing data. When our simulations more-so mimic situations that happen in the real world, we observe a stronger decline in model performance. This allows us to see more clearly the impact that missing data might have on underserved populations, whereas without this mechanism in place we might not have seen as much evidence of disparities.

One limitation of this work is the imprecise characterization of the actual causal pathways that occur in medical data. We remove clusters of related events as a proxy for these causal pathways, but in reality this may not accurately capture how missing data truly happens in practice. For example, a diagnosis might never occur without a certain abnormal lab test, and a prescription might never occur without a certain diagnosis. Thus, the causal pathway might be lab test → diagnosis → prescription. In all of our experiments, however, there is no direction in the pathways. We do not account for the fact that the probability of removing the lab test upstream in the causal pathway should yield a higher probability of removing the prescription downstream. Additionally, using the noisy OR-gate knowledge graph does not allow for the generation of failure probabilities relating events of the same type (such as two diagnosis events). Dependencies between events of the same type are still accounted for through their relationships with other events, but the method would be better if this were more explicit.

For future work, we might consider enlisting the help of clinicians that can identify general causal pathways that exist between medical events to further refine the knowledge graph and remove events in a more realistic way. We also might consider finding a way to map our EHRs events to existing databases that detail such relationships, such as the Google Health Knowledge Graph or the PennTURBO Knowledge Graph (Freedman et al., 2020). It is also of interest to account for missingness by developing novel imputation methods that can handle Level 1 EHRs and incorporate relatedness of medical concepts and events. There is a large body of literature on imputing missing data in EHRs, but little work on imputation models that can incorporate relatedness of medical concepts and events.

## Data Availability

All data produced in the present study are available upon reasonable request to the authors
https://physionet.org/content/mimiciii-demo/1.4/

## Acknowledgement

This work is partly supported by NIH grant R01-GM124111. The content is the responsibility of the authors and does not necessarily represent the views of NIH.

## Notes

### Competing Interest Statement

The authors have declared no competing interest.

### Author Declarations

This study involves only openly available medical data, which can be obtained from: https://physionet.org/content/mimiciii-demo/1.4/

